# Immunogenicity and Safety of a 14-valent pneumococcal polysaccharide conjugate vaccine (PNEUBEVAX 14^TM^) administered to 6-8 weeks old healthy Indian Infants: A single blind, randomized, active-controlled, Phase-III study

**DOI:** 10.1101/2023.12.21.23300357

**Authors:** Ramesh V Matur, Subhash Thuluva, Subbareddy Gunneri, Vijay Yerroju, Rammohan reddy Mogulla, Kamal Thammireddy, Piyush Paliwal, Niranjana S Mahantshetty, Mandyam Dhati Ravi, S. Prashanth, Savita Verma, Jai Prakash Narayan

**Affiliations:** Biological E Limited, 18/1&3, Azamabad, Hyderabad – 500 020, Telangana, India; KLES Dr. Prabhakar Kore Hospital & Medical Research Centre, Department of Pediatrics, Belgaum, Karnataka, India.; JSS Hospital, Department of Pediatrics, Mysore, Karnataka India; Cheluvamba Hospital, Mysore Medical College & Research Institute, Dept. of Paediatrics, Mysore, Karnataka, India; Pandit Bhagwat Dayal Sharma Post Graduate Institute of Medical Sciences & Hospital, Department of Pharmacology, Rothak, Haryana, India; Jawahar Lal Nehru Medical College, Kala Bagh, Rajasthan, India

**Keywords:** Infants, Pneumococcal disease, Conjugate vaccine, Serotype, Pneumococcal capsular polysaccharide, Opsonophagocytic activity

## Abstract

**Background:** Introduction of pneumococcal conjugate vaccines (PCV) reduced the cases of pneumococcal disease at global level. However, there is an increase in clinical and economic burden of PD from non-PCV serotypes, particularly in pediatric and elder population. In this study, immunogenicity and safety of the BE’s 14-valent PCV (PNEUBEVAX 14^TM^; BE-PCV-14) containing two additional epidemiologically important serotypes (22F and 33F) in comparison to PCV-13 was evaluated in infants.

**Methods:** This is a pivotal phase-3 single blind randomized active-controlled study conducted at 12 sites across India in 6-8 weeks old healthy infants in 6-10-14 weeks dosing schedule to assess immunogenic non-inferiority and safety of a candidate BE-PCV-14. In total, 1290 infants were equally randomized to receive either BE-PCV-14 or PCV-13. Solicited local reactions and systemic events, adverse events (AEs), serious AEs (SAEs) and medically attended AEs (MAAEs) were recorded. Immunogenicity was assessed by measuring anti-PnCPS IgG concentration and functional antibody titers by opsonophagocytic activity (OPA), one month after completing three dose schedule. Cross protection to serotype 6A offered by serotype 6B was also assessed in this study.

**Findings:** The safety profile of BE-PCV-14 was comparable to PCV-13 vaccine. Majority of reported AEs were mild in nature and no severe or serious AEs were reported. Primary immunogenicity objective was met for all 14 serotypes. For the twelve common serotypes non-inferiority to those 12 serotypes in PCV-13 was met. Additional serotypes in BE-PCV-14 (22F and 33F) also met NI criteria as defined by WHO TRS-977. A significant seroconversion, about 69% for serotype 6A was observed even though this antigen was not present in BE-PCV-14. This indicates that serotype 6B of BE-PCV-14 cross protects serotype 6A. BE-PCV-14 also elicited comparable serotype specific functional OPA immune responses to all the serotypes in PCV-13.

**Interpretations:** BE-PCV-14 was found to be safe and induced robust and functional serotype specific immune responses to all 14 serotypes. All serotype-specific IgG responses were comparable to those in PCV-13. These findings suggest that BE-PCV-14 can be safely administered to infants and achieve protection against pneumococcal disease caused by serotypes covered in the vaccine.

The study was prospectively registered with clinical trial registry of India-CTRI/2020/02/023129

## INTRODUCTION

*Streptococcus pneumoniae* is a gram positive bacterium, causing morbidity and mortality globally in children under 5 years, adults above 50 years and in individuals with comorbidities. It manifests a variety of invasive or noninvasive diseases such as pneumonia, otitis media, nasosinusitis, bacteremia and meningitis ^1,2^ Most of the deaths are caused by invasive pneumococcal disease (IPD) with reported >300000 deaths worldwide in children under 5 years every year due to pneumonia or meningitis ^3^ In developing countries, case fatality rates may be as high as 50% for meningitis and up to 20% for septicemia. As per a global population-based model in 2015 and as per WHO data, case fatality rates under 5 years’ is the highest in India followed by Pakistan, Bangladesh, Sri Lanka and Indonesia with reported incidences over 200/100,000 population ^4,5^ Although there are more than 95 identified serotypes of *S pneumoniae*, few serotypes are responsible for causing the disease ^6^. In addition, in more than 30% of cases, pneumococcal bacteria are resistant to one or more antibiotics ^7^.

A 23-valent, unconjugated pneumococcal polysaccharide vaccine (PPSV23; Pneumovax® 23, Merck and Co., Inc., NJ) provides only transient protection as it elicits T-cell independent immune responses^8^. Pneumococcal polysaccharide-protein conjugate vaccines (PCVs) 7- and 13-valent (PCV-7; Prevenar 7^®^; PCV-13; Prevenar 13^®^; Pfizer, New York), were able to induce T-cell dependent immune response against respective serotypes and generate robust functional antibody responses providing long-term protection against IPD across different age groups ^9–11^. Introduction of PCVs substantially reduced the burden of IPD worldwide in all age groups ^12^. Despite these reductions, one-fourth to one-third of the *S. pneumoniae* serotypes causing IPD are not covered by currently available PCVs and these serotypes bring about a significant unmet medical need ^13–15^. A study by Balsells, et al. ^16^ reported that 42.2% of childhood IPD cases were caused by non-vaccine serotypes. So, it is important to include additional serotypes to minimize the burden of IPD. Biological E Limited developed a PCV covering 14 different serotypes (PNEUBEVAX 14^TM^) including 2 serotypes (22F and 33F) that are not included in PCV-13. These two serotypes were reported to be among top 11 predominant non-vaccine serotypes, which accounted for 22% of invasive pneumococcal disease cases in India under-five years ^17,18^. A recent study ^19^, also reported that 22F and 33F as the most prevalent non-PCV13 serotypes causing IPD ^20^. Moreover, these two serotypes are among the first to increase in replacement disease after introduction of PCV-7 and PCV-13 vaccines ^19, 20^. Accessibility to PCVs with additional serotypes can be increased when the vaccines are developed in a country such as India that is among countries with the major disease burden.

Safety and immunogenicity of BE-PCV-14 was established in phase-1 and phase-2 studies in healthy adults and toddlers respectively (CTRI/2017/06/008759 and CTRI/2017/11/010387). Here we report the immunogenicity and safety of BE-PCV-14 in 6-8 weeks old healthy infants (n=1290) in 6-10-14 weeks dosing schedule and PCV-13 was used as a comparator to demonstrate the immunogenic non-inferiority.

## METHODS

### Study Design and Participants

The phase-3, two arm, single blind, randomized, active-controlled study was conducted between February 2020 and March 2022 in India across 12 sites. Study was designed to evaluate the immunogenic non-inferiority and safety of a BE-PCV-14 administered to 6-8 weeks old healthy infants in 6-10-14 weeks dosing schedule. Participants were equally randomized (1:1) to receive either BE-PCV-14 (test) or PCV-13 (Prevenar 13^®^, comparator). In total, 1290 subjects were enrolled into the study (n=645 in BE-PCV-14 arm and n=645 in PCV-13 arm). A dose of 0.5mL of the study vaccine (BE-PCV-14 or PCV-13) was administered intramuscularly in a three dose vaccination schedule (3+0) with a 4-weeks interval.

All enrolled subjects (N=1290) were healthy infants with ≥3300 grams weight at the time of screening. None of the subjects received any licensed or investigational pneumococcal vaccine prior to enrollment. Other permitted vaccines included single dose of BCG, hepatitis B, DTwP-HepB-Hib (liquid pentavalent), polio and rotavirus vaccines. Informed consent was obtained from subjects’ parent(s) prior to performing any study specific procedure. Exclusion criteria used were evidence of previous *S. pneumoniae* infection, use of any investigational or non-registered product (drug or vaccine) -30 days prior to study vaccine administration, history of any neurological disorders, meningitis, or seizures, family history of congenital or hereditary immunodeficiency, history of allergic disease or history of a serious reaction to any prior vaccination or known hypersensitivity likely to be exacerbated by any component of the study vaccines. Complete list of eligibility criteria provided as supplementary information.

The study was conducted in accordance with the principles defined in the Declaration of Helsinki, International Conference on Harmonization guidelines (Good Clinical Practices), and the new drug and clinical trial rules, 2019 ^21^. The investigational review board or ethics committee at each study site approved the protocol.

### Randomization and masking

Eligible subjects were randomized to receive either BE-PCV-14 or PCV-13 in 1:1 ratio and randomization numbers were assigned using an interactive web-based randomization system (IWRS). Randomization sequence was generated using SAS software (SAS Institute Inc, 100 SAS Campus Drive, Cary, NC 27513-2414, USA).

### Procedure

Screening, enrolment, and first dose of primary vaccination was done when infants were 6-8 weeks old (visit-1; day 0). Second and third doses of primary vaccination (visit-2; day 28 and visit-3; day 56) and a final follow up immunogenicity visit (visit-4, day 86) took place at four weeks’ interval.

An additional time window of +7 days for the second, third, and the fourth visit was permitted to ensure subject’s visit compliance.

Each 0.5mL of single human dose of BE-PCV-14 (PNEUBEVAX 14^TM^) contained 3μg of serotype 1 and 2.2μg of each of serotypes 3, 4, 5, 7F, 9V, 14, 18C, 19A, 19F, 22F, 23F and 33F and 4.4μg of serotype 6B polysaccharides conjugated to about 35 μg of non-toxic diphtheria toxin cross-reacting material 197 (CRM_197_) protein and adsorbed onto ≤0.75 mg of aluminium phosphate (Al+++). The 13-valent Pneumococcal polysaccharide conjugate vaccine “PCV-13” (control vaccine) contained 2.2μg of each of serotypes 1, 3, 4, 5, 6A, 7F, 9V, 14, 18C, 19A, 19F, 23F saccharides, and 4.4μg of 6B saccharide, 34μg CRM_197_ carrier protein, and 125μg aluminum as aluminum phosphate adjuvant. Both vaccines were administered in the anterolateral aspect of thigh.

Approximately 5.0 mL of blood was collected at visit-1(day 0) and again at visit-4 (day 86). Anti-PnCPS IgG antibody concentration estimation against each of the 14 vaccine serotypes was performed as per the WHO reference ELISA assay ^22^ with a minor modification. Cell wall polysaccharide (CWPS) multi was used instead of CWPS and 22F PnPS for pre-absorption to neutralize non-specific antibodies and other common contaminants present in the PnPS coating antigens. Functional antibody responses against all 14 serotypes were assessed by opsonophagocytic activity (OPA) assay, which was performed at WHO pneumococcal serology reference laboratory (UCL Great Ormond Street, Institute of Child Health, Immuno-biology Section, United Kingdom).

Each vaccinated infant was observed for at least 60 minutes after vaccination to evaluate and treat for any immediate adverse reactions. At each vaccination visit, a diary card was provided to the subject’s parent(s)/LAR[s] to record any solicited local/general AE occurring after vaccination for 7 consecutive days (day 0 - day 6). List of solicited local and systemic AEs are provided as supplementary information.

Unsolicited AEs, serious adverse events (SAEs), medically attended adverse events (MAAEs) were recorded until the end of the study. Severity of all AEs was graded as per the common terminology criteria for adverse events (CTCAE V5.0) or division of AIDS (DAIDS) table version 2.0. Brighton collaboration scale was taken for severity grading of fever. All AEs were assessed for relatedness to study vaccine by the investigator.

### Outcomes

Primary outcome of the study was to demonstrate serotype specific immunogenic non-inferiority (NI) of BE-PCV-14 to licensed PCV-13 vaccine one month after the completion of primary vaccination series against 12 common serotypes (1, 3, 4, 5, 6B, 7F, 9V, 14, 18C, 19A, 19F, and 23F) in terms of number of subjects seroconverted and the ratio of IgG geometric mean concentrations (GMCs). Immunogenic NI for new serotypes 22F and 33F in BE-PCV-14 was demonstrated by comparing immunoglobulin G (IgG) concentrations induced by serotypes 22F and 33F of BE-PCV-14 with one of the PCV-13 serotypes that showed the lowest sero-response. Sero-response or seroconversion rate was defined as proportion of vaccinated infants with serotype specific IgG antibody concentrations of at least ≥0.35μg/ml ^23^.

One of the secondary objectives of the study was to assess the immune response in terms of serotype specific opsonophagocytic activity (OPA) in a randomly chosen subset of subjects from the BE-PCV-14 group compared with an equal number of subjects randomly chosen from PCV-13 vaccinated group. Other secondary outcome was to evaluate the safety of BE-PCV-14 in comparison with PCV-13 measured by the incidence rates of reactogenicity events (local reactions and systemic events) and adverse events (AEs).

### Statistical Analyses

#### Sample size determination

Sample size estimation was based on the 2-sided 95% confidence interval (CI) for the difference in proportion of subjects achieving a serotype-specific pneumococcal IgG concentration ≥0.35μg/mL in each vaccine group. After study completion, a two-sided 95% confidence interval for the true difference between the two proportions was constructed. The lower bound of the confidence interval was to lie entirely on the positive side of the -10% points margin to demonstrate non-inferiority.

Data from the phase-II safety and immunogenicity study in 12 to 23 months old children conducted in India (CTRI/2017/11/010387) was used to support sample size estimation. The power was computed for an anticipated test vaccine group proportion of 92.33% within the non-inferiority margin set. The significance level of the test was targeted at 0.025. Accordingly, sample sizes of 581 in each group was estimated to achieve 90% power and to detect a non-inferiority margin difference between the group proportions of -0.10. An anticipated, not more than 10% dropout rate was built-in to the sample size. Therefore, the overall sample size required was 645 infants per group to obtain 581 evaluable infants per group at the end of the study.

### Statistical analyses

All demographic and baseline characteristics of both the groups were presented using intend-to treat (ITT) population, defined as all subjects randomized into the study. The data was analyzed by summary statistics. All subjects who received at least one dose of the study vaccine were included in the safety analysis and the datawas summarized descriptively. For continuous variables *n, mean, standard deviation, median* and range (minimum, maximum) have been presented. For categorical data, frequencies were computed. All reported adverse events during the entire study period were summarized by calculating frequencies and listed per subject including severity and relationship (causality).

The number and percentage of subjects with AEs were presented by system organ class and preferred term. All medically attended AEs reported during the study were listed and analyzed for expectedness and causality. Two-sided 95% exact confidence intervals were calculated for all the occurrence rates of reported AEs and SAEs during the study. All AEs were coded using the Medical Dictionary for Regulatory Activities (MedDRA^TM^; version 25.1) coding dictionary and concomitant medications have been coded using the World Health Organization (WHO) Drug Dictionary, March 2022.

Immunogenicity assessment was based on the per-protocol population defined as the vaccinated subjects whose blood samples were available for immunogenicity analysis at both baseline and day 86 (visit-4). Non-inferiority of serotype specific immune responses induced by BV-PCV-14 against PCV-13 was assessed by one of the below two criteria as per the WHO defined criteria^23^.

1. By measuring difference in proportion of subjects seroconverted to achieve serotype specific anti-PnCPS IgG concentration ≥ 0.35 μg/ml between BE-PCV-14 and PCV-13. The lower limit of 95% CI for the difference in proportion subjects seroconverted between BE-PCV-14 and PCV-13 should be above the minus 10% points.
2. By assessing the IgG GMC ratio (BE-PCV-14/PCV-13) along with its two sided 95% confidence interval. Non-inferiority was considered achieved if the ratio of lower limit of two sided 95% CI (test /control group) was above the pre-specified margin of 0.5.

For two new serotypes 22F and 33F which were not part of PCV-13, non-inferiority was assessed by comparing with any serotype in the PCV-13 group that achieved the lowest percentage of ≥0.35μg/ml IgG GMC. For serotype 6A, which was not in BE-PCV-14 but present in PCV-13, a ≥2-fold increase in the 6A specific geometric mean concentrations (GMCs) above pre-vaccination levels was assessed as an exploratory outcome. Proportion of subjects achieving ≥4-fold rise in anti-PnCPS IgG antibody concentrations from baseline and geometric mean fold rise (GMFR) along with their 2-sided 95% CI against each of the vaccine serotype was calculated.

As a secondary outcome, proportion of subjects achieving a serotype-specific OPA titre≥1:8 post vaccination was assessed along with 2-sided 95% confidence. Serotype specific OPA geometric mean titres (GMT) along with their ratios and GMFR against 12 common serotypes in both vaccine groups was evaluated.

A reverse cumulative distribution (RCD) curve of anti-PnCPS antibody concentrations and OPA GMT titers by serotype of BE-PCV-14 and PCV-13 were plotted for all common vaccine serotypes in both groups.

## RESULTS

A total of 1290 subjects were screened and randomly assigned to receive either BE-PCV-14(n=645) or PCV-13 (n=645) study vaccine. Out of 1290 subjects, 1267 (98.2%) subjects (641 in BE-PCV-14 arm and 626 in PCV-13 arm) completed all the visit specific procedures and were included in the immunogenicity (anti-PnCPS IgG antibody concentration) analysis. In total, 22 (1.7%) subjects were dropouts. Subject disposition with reason for dropout from the study was presented in Figure 1. Safety analyses were performed in all enrolled subjects (n=1290) whereas immunogenicity (per-protocol population) was completed in 1267 subjects. One subject was excluded from per-protocol population due to insufficient serum sample and the rest (n=22) were dropouts. Demographics of intention-to-treat population were presented in Table 1. Age of the subjects and male to female ratio was comparable in both BE-PCV-14 and PCV-13 arms. Summary of other eligible vaccinations received by study subjects was presented in Supplementary Table 1.

**TABLE 1:**
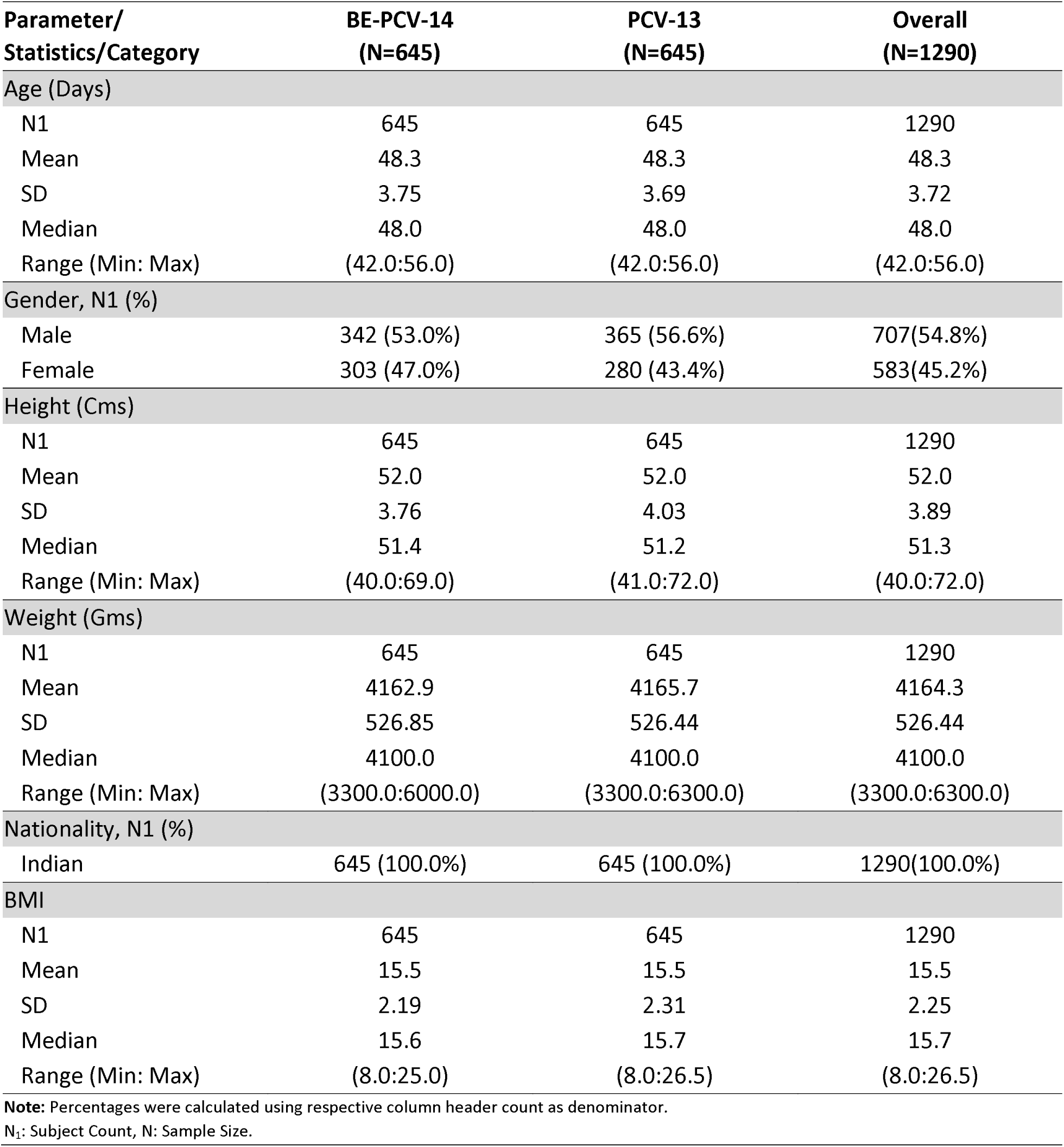
Demographic characteristics of study participants

The proportion of subjects with anti-PnCPS IgG antibody concentrations equal to or above the protective threshold ≥0.35µg/mL (primary outcome) at day 86 in BE-PCV-14 (n=641) group was in the range of 76.3% (serotype6B) to 99.7% (serotype 14) and in the PCV-13 (n=626) group, the response rate was in the range of 77% (serotype 3) to 100% (serotype14) for 12 common serotypes. The lower limit of 95% CI for the difference in proportion of subjects seroconverted between BE-PCV-14 and PCV-13 was above the minus 10% points for all the 12 common serotypes (serotype 1, 3, 4, 5, 6B, 7F, 9V, 14, 18C, 19A, 19F and 23F) and all of them met pre-defined NI criteria. The distributions in serotype-specific IgG concentrations for 12 common serotypes in both vaccine groups were similar, as shown by the reverse cumulative distribution curves (Figure 3).

Seroresponse rates for serotype 22F (94.1%) and 33F (73.2%) that are present only in BE-PCV-14 group were compared with the lowest performing serotype 3 (77.0%) in PCV-13 group. Defined NI criteria was met for both serotypes 22F and 33F (Figure 2a).

**Figure 2a:**
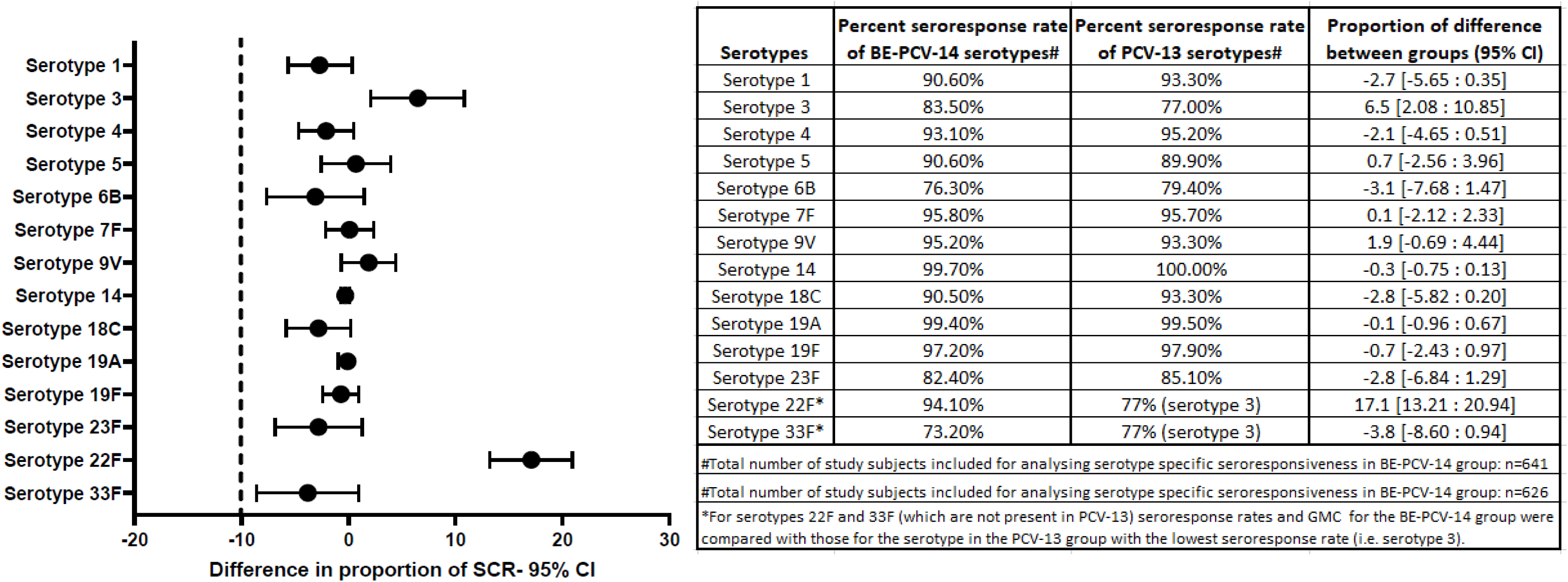
Difference in proportion of seroconversion rates with 95% confidence intervals

For IgG geometric mean concentrations, NI criteria was achieved for all 12 common serotypes and for additional 22F and 33F serotypes (Figure 2b). The lower bound of the 95% CI for GMC ratio was in the range of 0.67 (serotype 1) to 1.10 (serotype 3) for 12 common serotypes present in BE-PCV-14 and PCV-13. For serotype 3 and serotype 14, lower bound 95% CI was 1.10 and 1.01 respectively, indicating that BE-PCV-14 induces strong immune response compared to PCV-13. Serotypes 22F and 33F, which are not present in PCV-13, the lower bound of the 95% CI for GMC ratio was 5.26 and 1.82 respectively (Figure 2b), when compared to the lowest performing serotype in PCV-13 (serotype 3).

**Figure 2b:**
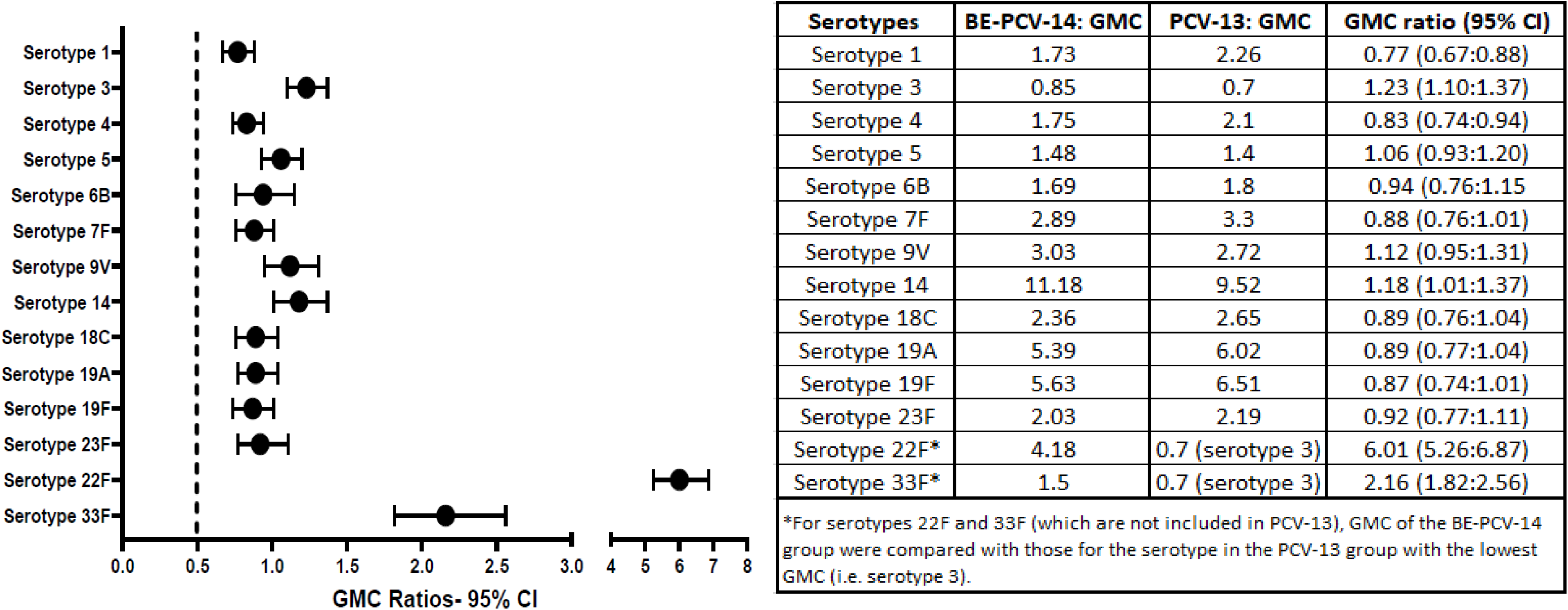
Geometric Mean Concentration ratios with 95% confidence intervals

**Figure 3:**
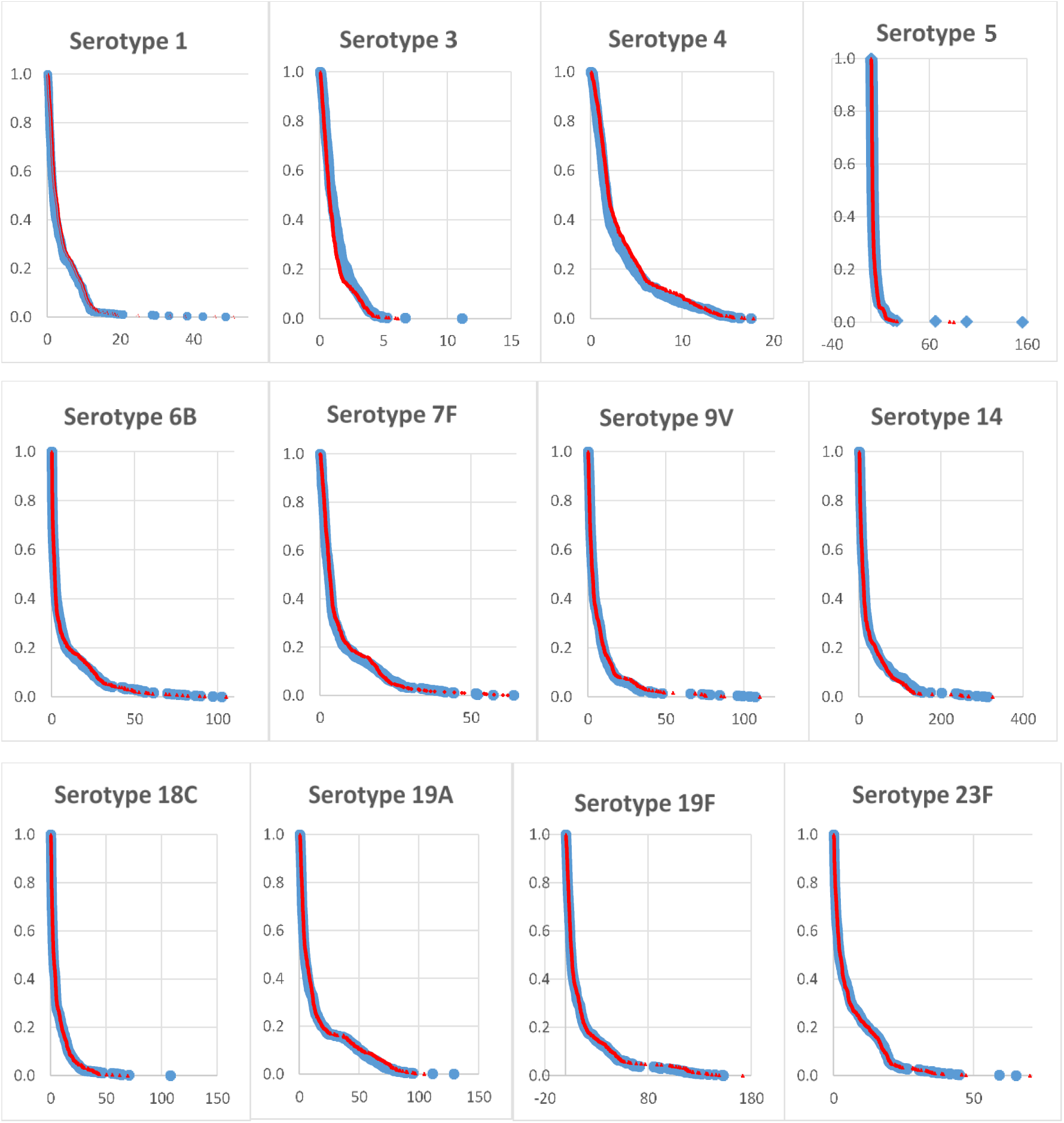
Reverse cumulative distribution (RCD) curves of anti-PnCPS IgG antibody concentrations for 12 common serotypes between BE-PCV-14 and PCV-13.

The proportion of subjects achieving a serotype-specific OPA titer ≥LLOQ (≥1:8) at day 86 were in the range of 63.64% (serotype-1) to 98.43% (serotype-22F) in BE-PCV-14 group and 64.43% (serotype-1) to 98.46% (serotype-7F and serotype-23F) in PCV-13 group. Overall, the OPA GMTs were comparable among all common serotypes in both vaccinated groups (Table 2).

**Table 2:**
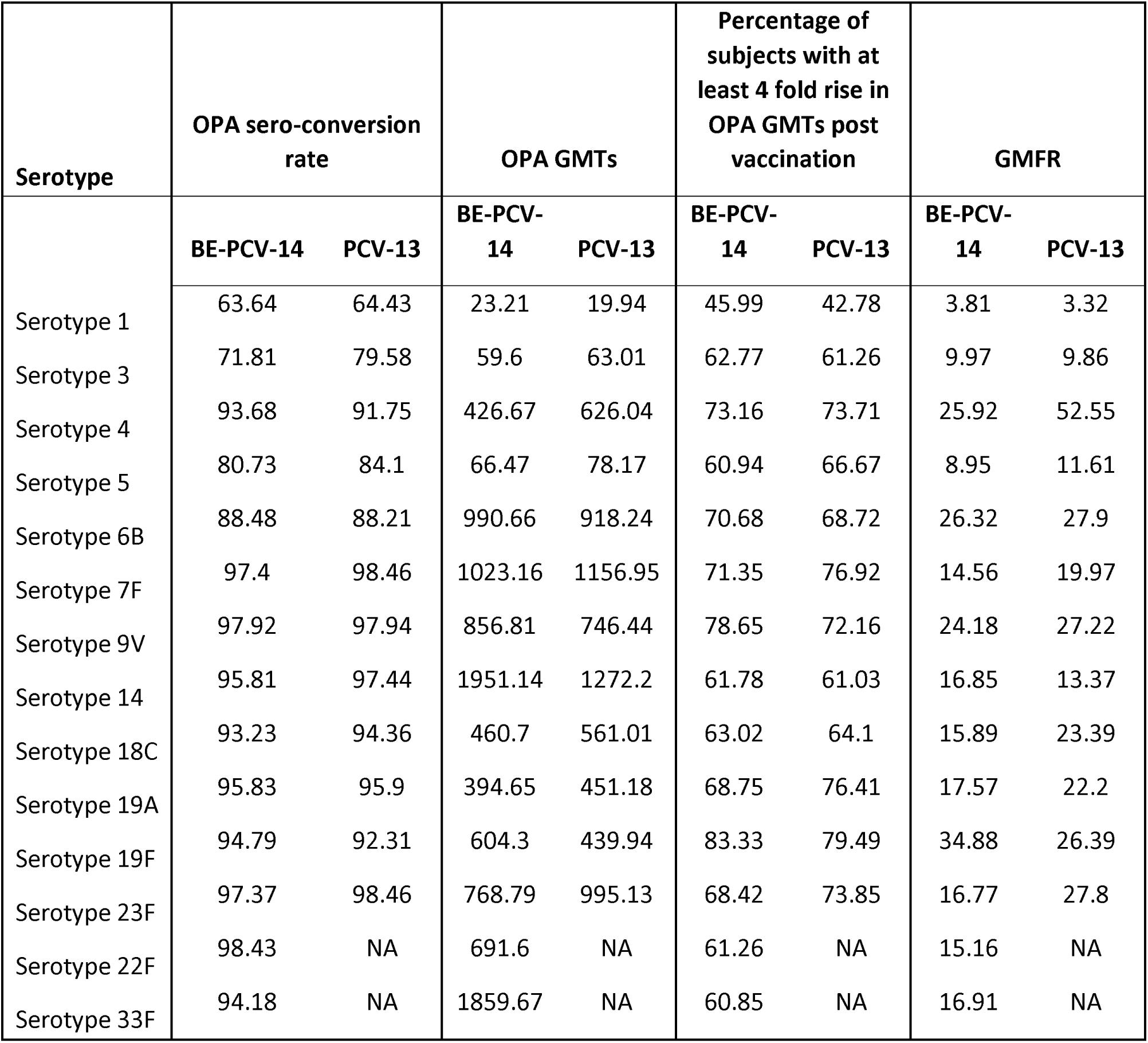
Summary of serotype-specific OPA titer ≥LLOQ (≥1:8) in terms of sero-conversion rates, GMTs and fold rise.

The additional serotypes 22F and 33F in BE-PCV-14 vaccine had 15.16 and 16.91-fold rise in OPA GMTs respectively at day 86 compared to pre-vaccination levels, signifying that both the additional serotypes can induce high levels of protective functional antibodies in vaccinated infants (Table 2). The proportion of subjects achieving ≥4-fold increase in anti-PnCPS IgG antibody concentrations at day 86 from baseline is presented in Table 3.

**Table 3:**
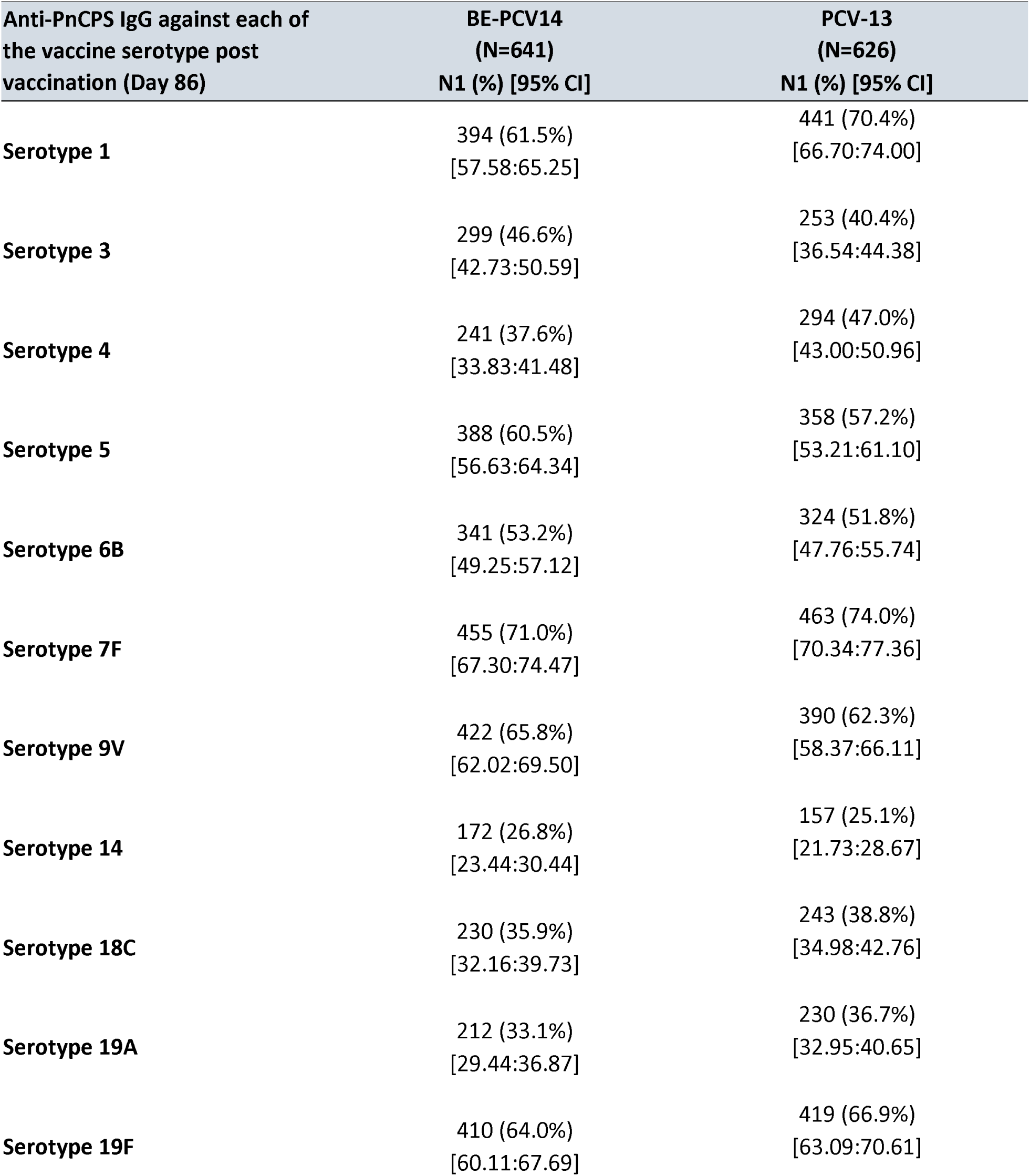
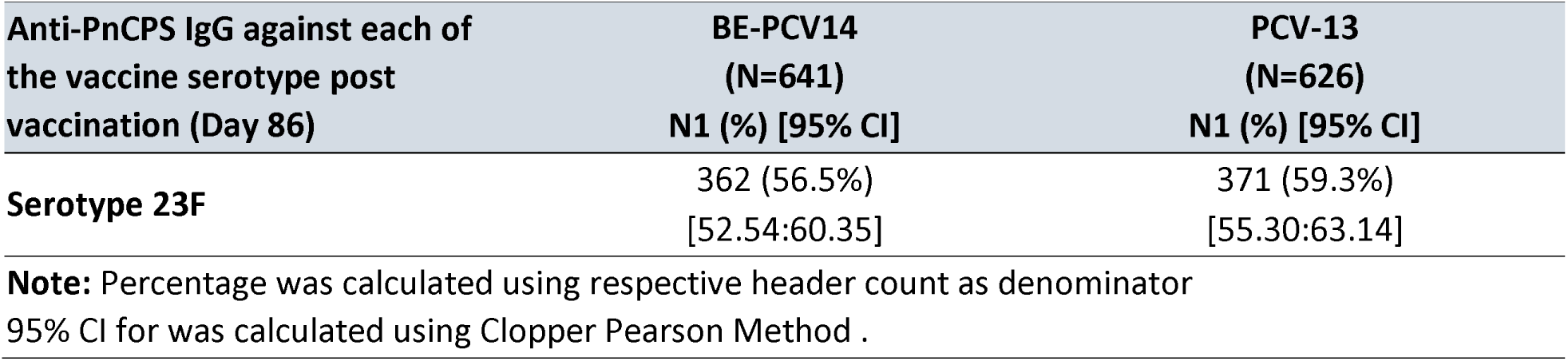
Proportion of subjects achieving **≥**4-fold increase in anti-PnCPS IgG antibody

BE-PCV-14 vaccine has serotype 6B and no serotype 6A. In this study, we tested the cross protection against serotype 6A, assessed by a ≥2-fold increase in the 6A specific IgG antibody concentrations and the OPA antibody titers above the pre-vaccination levels. There was 2.4-fold (pre-vaccination GMC: 0.45 and post-vaccination GMC: 1.08) and 7.64-fold (OPA GMT were 16.59 and 134.50 for pre-and post-vaccination, respectively) increase in serotype 6A specific IgG and OPA antibody titers respectively (Table 4). Our data on serotype 6A specific 69.7% seroconversion and 2.4-fold rise in GMC in BE-PCV-14 vaccinated infants along with functional antibody titers (OPA) GMFR of 7.64 fold strongly suggests that the serotype 6B confers significant immunological cross protection against serotype 6A.

**Table 4:** Serotype-6A specific IgG and OPA immune responses:

| **ST-6A**<br>**IgG**<br>**GMC**<br>**(N=1290)** | **Pre-Vac** | **Post-Vac** | **GMFR** | **ST-6A**<br>**OPA**<br>**GMT**<br>**(N=386)** | **Pre-Vac** | **Post-Vac** | **GMFR** |
| --- | --- | --- | --- | --- | --- | --- | --- |
| **BE-PCV-**<br>**14**<br>**(n=645)** | 0.448 | 1.075 | **2.4** | **BE-**<br>**PCV14**<br>**(n=192)** | 16.59 | 134.50 | **7.64** |
| **PCV-13**<br>**(n=645)** | 0.446 | 1.906 | **4.27** | **PCV-13**<br>**(n=194)** | 19.20 | 690.63 | **37.35** |

The current study demonstrated that BE’s 14-valent pneumococcal polysaccharide conjugate vaccine, administered in a 3-dose schedule (6-10-14 weeks), was safe, immunogenic and non-inferior to PCV-13 for the 12 common serotypes.

Among all vaccinated infants (n=1290), 175/645 (27.1%) subjects and 178/645 (27.6%) subjects reported at least one adverse event in BE-PCV-14 and PCV-13 groups respectively. All the reported adverse events were generally mild and only very few cases were moderate in their intensity. Majority of the reported adverse events were solicited in nature and related to the study vaccine. Overview of the reported AEs including solicited and unsolicited AEs were presented in Table 5. Summary of AEs by system organ class (SOC) and preferred term (PT) were shown in Table 6. No severe or serious AEs were reported in either of the vaccine groups. Overview of AEs by severity grade and causality to the study vaccine were presented in Table 7. Summary of local, solicited systemic and unsolicited AEs by SOC and PT were presented in Supplementary Table 2, 3 and 4. No clinically significant changes overtime were noted in the vital signs. The AEs observed and physical examination results did not indicate any safety issues of concern. Overall, the safety profile of BE-PCV-14 was comparable to the control vaccine PCV-13 in terms of AE rates, related AE rates and medically attended AEs and found to be safe and well tolerated.

**Table 5:** Overview of AEs-solicited and unsolicited AEs

| Category<br>N1 (%) [95% CI] n | BE-PCV-14<br>(N=645) | PCV-13<br>(N=645) |
| --- | --- | --- |
| Overall AEs | 175 (27.1%)<br>[23.73:30.74] 355 | 178 (27.6%)<br>[24.18:31.22] 353 |
| Serious AEs | 0 (0.0%) [NE] 0 | 0 (0.0%) [NE] 0 |
| Severe AEs | 0 (0.0%) [NE] 0 | 0 (0.0%) [NE] 0 |
| Local AEs | 137 (21.2%)<br>[18.14:24.60] 235 | 144 (22.3%)<br>[19.17:25.74] 251 |
| Systemic AEs | 70 (10.9%)<br>[8.56:13.51] 118 | 63 (9.8%)<br>[7.59:12.32] 101 |
| Medically Attended AEs | 0 (0.0%) [NE] 0 | 0 (0.0%) [NE] 0 |
| AEs within 60 minutes postvac | 4 (0.6%)<br>[0.17:1.58] 6 | 4 (0.6%)<br>[0.17:1.58] 5 |
| AEs within 7 days postvac | 174 (27.0%)<br>[23.59:30.58] 354 | 178 (27.6%)<br>[24.18:31.22] 352 |
| AEs between Day 0 to Day 28 | 115 (17.8%)<br>[14.95:21.01] 207 | 109 (16.9%)<br>[14.09:20.02] 190 |
| AEs between Day 28 to Day 56 | 61 (9.5%)<br>[7.31:11.98] 93 | 73 (11.3%)<br>[8.98:14.02] 110 |
| AEs between Day 56 to Day 86 | 46 (7.1%)<br>[5.27:9.40] 55 | 41 (6.4%)<br>[4.60:8.52] 53 |
| Any Solicited AEs | 174 (27.0%)<br>[23.59:30.58] 353 | 178 (27.6%)<br>[24.18:31.22] 352 |
| Any Unsolicited AEs | 2 (0.3%)<br>[0.04:1.12] 2 | 1(0.2%)[0.00:0.86]1 |

**Note:** Percentages were calculated using respective column header group count as denominator.
95% CI was calculated by Clopper-Pearson Method.
N<sub>1</sub>: Subject Count, N: Subject Count, N: Sample Size, n: Event Count

**Table 6:** Summary of AEs by SOC and PT

| SOC/PT,<br>N1 (%) [95% CI] n | BE-PCV-14<br>(N=645) | PCV-13<br>(N=645) |
| --- | --- | --- |
| Overall | 175 (27.1%) [23.73:30.74] 355 | 178 (27.6%) [24.18:31.22] 353 |
| Gastrointestinal disorders | 9 (1.4%) [0.64:2.63] 12 | 5 (0.8%) [0.25:1.80] 8 |
| Abdominal pain | 1 (0.2%) [0.00:0.86] 1 | 0 (0.0%) [NE] 0 |
| Diarrhoea | 6 (0.9%) [0.34:2.01] 8 | 2 (0.3%) [0.04:1.12] 3 |
| Vomiting | 3 (0.5%) [0.10:1.35] 3 | 4 (0.6%) [0.17:1.58] 5 |
| General disorders and<br>administration site conditions | 169 (26.2%) [22.85:29.78] 329 | 173 (26.8%) [23.44:30.42] 333 |
| Injection site erythema | 33 (5.1%) [3.55:7.11] 38 | 36 (5.6%) [3.94:7.64] 44 |
| Injection site induration | 9 (1.4%) [0.64:2.63] 13 | 13 (2.0%) [1.08:3.42] 18 |
| Injection site pain | 93 (14.4%) [11.80:17.37] 133 | 98 (15.2%) [12.51:18.20] 131 |
| Injection site swelling | 44 (6.8%) [5.00:9.05] 51 | 48 (7.4%) [5.54:9.75] 58 |
| Irritability Postvaccinal | 25 (3.9%) [2.52:5.67] 41 | 21 (3.3%) [2.03:4.93] 31 |
| Pyrexia | 42 (6.5%) [4.73:8.70] 53 | 40 (6.2%) [4.47:8.35] 51 |
| Metabolism and nutrition<br>disorders | 8 (1.2%) [0.54:2.43] 9 | 6 (0.9%) [0.34:2.01] 10 |
| Decreased appetite | 8 (1.2%) [0.54:2.43] 9 | 6 (0.9%) [0.34:2.01] 10 |
| Nervous system disorders | 2 (0.3%) [0.04:1.12] 4 | 1 (0.2%) [0.00:0.86] 2 |
| Somnolence | 2 (0.3%) [0.04:1.12] 4 | 1 (0.2%) [0.00:0.86] 2 |
| Respiratory, thoracic and<br>mediastinal disorders | 1 (0.2%) [0.00:0.86] 1 | 0 (0.0%) [NE] 0 |
| Cough | 1 (0.2%) [0.00:0.86] 1 | 0 (0.0%) [NE] 0 |

**Note:** Percentages were calculated using column header group count as denominator.
95% CI were calculated by Clopper -Pearson Method.
N1: Subject Count, N: Sample Size, n: Event Count

**Table 7:**
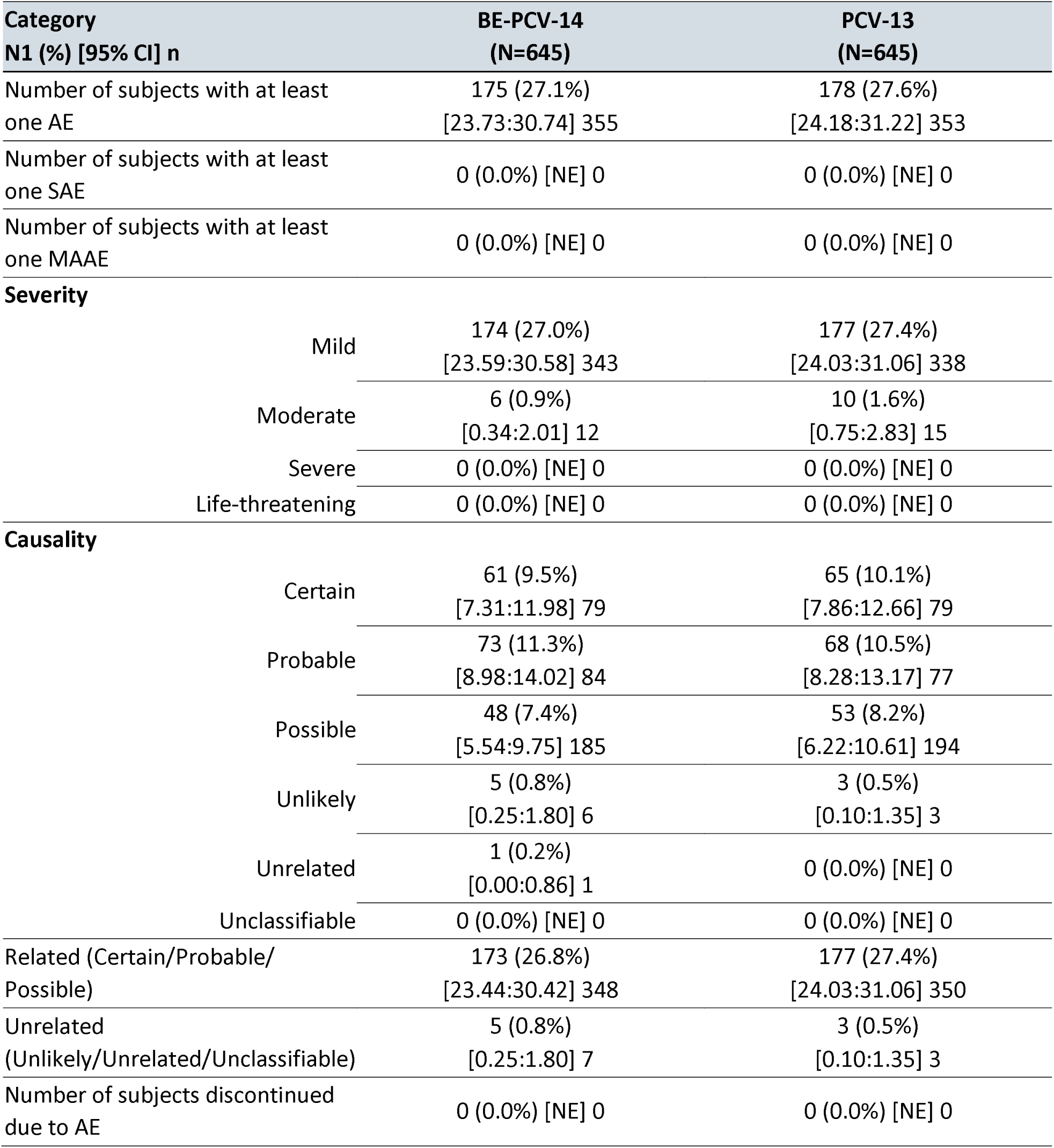
Overview AEs by Severity & Causality

| Category<br>N1 (%) [95% CI] n |  | BE-PCV-14<br>(N=645) | PCV-13<br>(N=645) |
| --- | --- | --- | --- |
| Number of subjects with at least one AE |  | 175 (27.1%)<br>[23.73:30.74] 355 | 178 (27.6%)<br>[24.18:31.22] 353 |
| Number of subjects with at least one SAE |  | 0 (0.0%) [NE] 0 | 0 (0.0%) [NE] 0 |
| Number of subjects with at least one MAAE |  | 0 (0.0%) [NE] 0 | 0 (0.0%) [NE] 0 |
| **Severity** |  |  |  |
|  | Mild | 174 (27.0%)<br>[23.59:30.58] 343 | 177 (27.4%)<br>[24.03:31.06] 338 |
|  | Moderate | 6 (0.9%)<br>[0.34:2.01] 12 | 10 (1.6%)<br>[0.75:2.83] 15 |
|  | Severe | 0 (0.0%) [NE] 0 | 0 (0.0%) [NE] 0 |
|  | Life-threatening | 0 (0.0%) [NE] 0 | 0 (0.0%) [NE] 0 |
| **Causality** |  |  |  |
|  | Certain | 61 (9.5%)<br>[7.31:11.98] 79 | 65 (10.1%)<br>[7.86:12.66] 79 |
|  | Probable | 73 (11.3%)<br>[8.98:14.02] 84 | 68 (10.5%)<br>[8.28:13.17] 77 |
|  | Possible | 48 (7.4%)<br>[5.54:9.75] 185 | 53 (8.2%)<br>[6.22:10.61] 194 |
|  | Unlikely | 5 (0.8%)<br>[0.25:1.80] 6 | 3 (0.5%)<br>[0.10:1.35] 3 |
|  | Unrelated | 1 (0.2%)<br>[0.00:0.86] 1 | 0 (0.0%) [NE] 0 |
|  | Unclassifiable | 0 (0.0%) [NE] 0 | 0 (0.0%) [NE] 0 |
| Related (Certain/Probable/Possible) |  | 173 (26.8%)<br>[23.44:30.42] 348 | 177 (27.4%)<br>[24.03:31.06] 350 |
| Unrelated (Unlikely/Unrelated/Unclassifiable) |  | 5 (0.8%)<br>[0.25:1.80] 7 | 3 (0.5%)<br>[0.10:1.35] 3 |
| Number of subjects discontinued due to AE |  | 0 (0.0%) [NE] 0 | 0 (0.0%) [NE] 0 |

## DISCUSSION

In this phase-3 trial, immunogenic non-inferiority, safety, and reactogenicity of a candidate BE-PCV-14 vaccine administered to 6-8 weeks old healthy infants in 6-10-14 weeks dosing schedule was evaluated. Robust and functional immune responses were elicited by BE-PCV-14 one month after the third vaccination dose. Primary basis for NI comparison and licensure decisions were based on reaching IgG concentrations ≥0.35µg/ml for all vaccine specific serotypes in the new vaccine and the licensed comparator. In accordance with WHO TRS 977 guidelines ^23^, all 12 common serotypes in BE-PCV-14 met the non-inferiority criteria with those serotypes in PCV-13 in terms of both seroconversion rates (percentage of subjects with IgG ≥0.35µg/ml) and GMC ratios (≥0.5). Both BE-PCV-14 and PCV-13 generated comparable functional immune responses (OPA titres), similar to IgG response.

In the current study, we evaluated the cross-protection offered by serotype 6B against serotype 6A in BE-PCV-14 vaccinated group. Serotype 6A and 6B have high structural similarity of polysaccharide ^24,25^. Therefore, it was expected that the immune responses induced by serotype 6B would provide at least partial cross-protection against serotype 6A in humans. An earlier study reported that PCV-13 elicits cross-functional opsonophagocytic killing responses in humans to serotypes not directly covered by the vaccine ^26^. In this study, cross-protection was assessed by a ≥2-fold increase in the 6A specific IgG antibody concentrations and the OPA antibody titers above the pre-vaccination levels. We found 2.4-fold and 7.64-fold increase in IgG GMC and OPA titers respectively, against serotype 6A in BE-PCV-14 vaccinated subjects, suggesting that BE-PCV-14 vaccine can offer protection against serotype 6A even though this serotype antigen is not present in BE-PCV-14. The serotype 6A cross protective functional immune response elicited by BE-PCV-14 is notably robust and strong when compared to another 10-valent PCV as described elsewhere ^27^. This clearly indicates that serotype 6B in BE-PCV-14 affords significant cross protection to serotype 6A. Our results are consistent with previous studies, where it was shown that 23-valent polysaccharide vaccine lacking serotype 6A and carrying serotype 6B elicited cross-functional immune responses against serotype 6A ^28,29^.

Vaccine effectiveness (VE) against IPD was established in pediatric population for PCV-13 specific serotypes and it was found to be around 55% ^30^. Breakthrough IPD was mainly caused by serotype 3 and 19A in PCV-13 vaccinated children. Excluding serotype 3 and 19A, VE of PCV-13 against rest of the vaccine serotypes was reported to be 89% ^30^. BE-PCV-14 was non-inferior to PCV-13 for all shared serotypes and against 2 unique serotypes (22F and 33F) when compared against lowest performing serotype 3 from PCV-13. Importantly, 83.5% of BE-PCV-14 vaccinated children seroconverted against serotype 3 while 77% seroconverted to serotype 3 in PCV-13 group. Several studies worldwide have shown that substantial proportion of pneumococcal disease is caused by non-vaccine serotypes, predominantly by serotype 22F (13.4%) and 33F (21%) ^19,31,32^. Another 15 valent PCV carrying these two serotypes was shown to induce serotype 22F and 33F immune response when compared to PCV-13 ^33^. Similarly, the present study also demonstrates that BE-PCV-14 elicited serotype specific (22F and 33F) superior immune responses (Figure 2b).

BE-PCV-14 containing 12 common serotypes and 22F and 33F serotypes along with cross protection to serotype 6A can protect the susceptible population against pneumococcal disease and may also provide herd immunity.

One limitation of the current study is that it is conducted in only Indian infant population and need to conduct similar studies in other regions. However, based on the VE and post marketing experience with PCV-13 across different geographical regions, it is likely that BE-PCV-14 will have similar effectiveness across regions.

In conclusion, BE-PCV-14 was safe, well tolerated and showed comparable safety profile to that of PCV-13. BE-PCV-14 elicited immune responses non-inferior to PCV-13 in infants for all 12 common serotypes. Statistical NI was also met for two new serotypes 22F and 33F in BE-PCV-14 vaccine. BE-PCV-14 also induced cross protective antibodies against serotype 6A. These data support BE-PCV-14 protection against 14 pneumococcal serotypes and potential benefit against serotype 6A.

## Data Availability

All data produced in the present study are available upon reasonable request to the authors.

## Acknowledgement

We are thankful to all the study participants, the principal investigators, and the study staff at all the clinical sites. We are thankful to all the study participants, the principal investigators, and the study staff at all the clinical sites. The authors would like to thank Scientific Advisory Board (SAB) and the Management of Biological E Limited for their support and valuable guidance. All authors wish to express their appreciation and gratitude for all front-line healthcare workers. Development of this vaccine would not have been possible without the efforts of R&D team, manufacturing, quality control, quality assurance and regulatory teams at Biological E. We would also like to thank the members of the DSMB for safety monitoring of the study data. Authors would like to thank Dr. Pothakamuri Venkata Suneetha for her valuable support in drafting, reviewing and finalizing the manuscript, and Mr. Kalyan Kumar P, Mr. Raju Esanakarra & Mr. Naga Ganesh B for clinical site management and monitoring.

## Data Sharing Agreement

Study data presented in the manuscript can be made available upon request and addressed to the corresponding author Dr. Ramesh V Matur at his

