## Supplementary tables and figures for "Immunogenicity and Safety of a 14-valent pneumococcal polysaccharide conjugate vaccine (PNEUBEVAX 14^TM^) administered to 6-8 weeks old healthy Indian Infants: A single blind, randomized, active-controlled, Phase-III study"

**SUPPLEMENTARY TABLE 1**: **Summary of Vaccination Details - Safety Population (N=1290)**

| **Vaccination** | **BE-PCV14 (N=645)** | **Prevenar 13 (N=645)** | **Overall (N=1290)** |
| --- | --- | --- | --- |
| BCG | 645 (100.0%) | 645 (100.0%) | 1290 (100.0%) |
| Hepatitis-B | 643 (99.7%) | 644 (99.8%) | 1287 (99.8%) |
| Polio^#^ | 645 (100.0%) | 645 (100.0%) | 1290 (100.0%) |
| DTwP-HepB-Hib (Pentavalent) ^#^ | 645 (100.0%) | 645 (100.0%) | 1290 (100.0%) |
| **Note:** Percentage were calculated with respective column header count as denominator. N: Sample Size.  ^#^Concomitantly given along with PCV14 vaccine | | | |

**SUPPLEMENTARY TABLE 2: Summary of local, solicited systemic and unsolicited AEs by SOC and PT**

| **SOC/PT**  **N1 (%) [95% CI] n** | **BE-PCV14 (N=645)** | **Prevenar 13 (N=645)** |
| --- | --- | --- |
| Overall | 137 (21.2%) [18.14:24.60] 235 | 144 (22.3%) [19.17:25.74] 251 |
| General Disorders And Administration Site Conditions | 137 (21.2%) [18.14:24.60] 235 | 144 (22.3%) [19.17:25.74] 251 |
| Injection Site Erythema | 33 (5.1%) [3.55:7.11] 38 | 36 (5.6%) [3.94:7.64] 44 |
| Injection Site Induration | 9 (1.4%) [0.64:2.63] 13 | 13 (2.0%) [1.08:3.42] 18 |
| Injection Site Pain | 93 (14.4%) [11.80:17.37] 133 | 98 (15.2%) [12.51:18.20] 131 |
| Injection Site Swelling | 44 (6.8%) [5.00:9.05] 51 | 48 (7.4%) [5.54:9.75] 58 |
| **Note:** Percentages were calculated using column header count as denominator. 95% CI were calculated by Clopper -Pearson Method. N1: Subject Count, N: Sample Size, n: Event Count **General Note:** •   All AE's were represented as: Subject count (Percentage of subjects) [95% CI] Event Count. | | |

**SUPPLEMENTARY TABLE 3: Summary of Solicited Systemic AEs by SOC & PT**

| **Reported Term**  **N1 (%) [95% CI] n** | **BE-PCV14 (N=645)** | **Prevenar 13 (N=645)** |
| --- | --- | --- |
| Overall | 70 (10.9%) [8.56:13.51] 118 | 63 (9.8%) [7.59:12.32] 101 |
| Gastrointestinal Disorders | 8 (1.2%) [0.54:2.43] 11 | 5 (0.8%) [0.25:1.80] 8 |
| Diarrhoea | 6 (0.9%) [0.34:2.01] 8 | 2 (0.3%) [0.04:1.12] 3 |
| Vomiting | 3 (0.5%) [0.10:1.35] 3 | 4 (0.6%) [0.17:1.58] 5 |
| General Disorders And Administration Site Conditions | 64 (9.9%) [7.73:12.49] 94 | 58 (9.0%) [6.90:11.47] 81 |
| Irritability Postvaccinal | 25 (3.9%) [2.52:5.67] 41 | 21 (3.3%) [2.03:4.93] 31 |
| Pyrexia | 42 (6.5%) [4.73:8.70] 53 | 40 (6.2%) [4.47:8.35] 50 |
| Metabolism And Nutrition Disorders | 8 (1.2%) [0.54:2.43] 9 | 6 (0.9%) [0.34:2.01] 10 |
| Decreased Appetite | 8 (1.2%) [0.54:2.43] 9 | 6 (0.9%) [0.34:2.01] 10 |
| Nervous System Disorders | 2 (0.3%) [0.04:1.12] 4 | 1 (0.2%) [0.00:0.86] 2 |
| Somnolence | 2 (0.3%) [0.04:1.12] 4 | 1 (0.2%) [0.00:0.86] 2 |
| **Note:** Percentages were calculated using column header count as denominator. 95% CI were calculated by Clopper -Pearson Method. N1: Subject Count, N: Sample Size, n: Event Count **General Note:** • All AE's were represented as: Subject count (Percentage of subjects) [95% CI] Event Count. • Solicited Local and Systemic AEs were recored during 7 days (Day 0 – Day 6) after each dose. | | |

**SUPPLEMENTARY TABLE 4: Summary of Unsolicited Systemic AEs by SOC & PT**

| **SOC/PT, N1 (%) [95% CI] n** | **BE-PCV14 (N=645)** | **Prevenar 13 (N=645)** |
| --- | --- | --- |
| Overall | 2 (0.3%) [0.04:1.12] 2 | 1 (0.2%) [0.00:0.86] 1 |
| Gastrointestinal Disorders | 1 (0.2%) [0.00:0.86] 1 | 0 (0.0%) [NE] 0 |
| Abdominal Pain | 1 (0.2%) [0.00:0.86] 1 | 0 (0.0%) [NE] 0 |
| General Disorders And Administration Site Conditions | 0 (0.0%) [NE] 0 | 1 (0.2%) [0.00:0.86] 1 |
| Pyrexia | 0 (0.0%) [NE] 0 | 1 (0.2%) [0.00:0.86] 1 |
| Respiratory, Thoracic And Mediastinal Disorders | 1 (0.2%) [0.00:0.86] 1 | 0 (0.0%) [NE] 0 |
| Cough | 1 (0.2%) [0.00:0.86] 1 | 0 (0.0%) [NE] 0 |
| **Note:** Percentages were calculated using column header count as denominator. 95% CI was calculated by Clopper-Pearson Method. N_1_: Subject Count, N: Sample Size, n:Event Count. **General Note:** •  All AE's were represented as: Subject count (Percentage of subjects) [95% CI] Event Count. •  Unsolicited adverse event reported at any time, until 28 days after the 2nd dose. | | |
