## Supplementary information for "Immunogenicity and Safety of a 14-valent pneumococcal polysaccharide conjugate vaccine (PNEUBEVAX 14^TM^) administered to 6-8 weeks old healthy Indian Infants: A single blind, randomized, active-controlled, Phase-III study"

Subjects were enrolled in the study based on the following inclusion criteria.

1. Healthy pneumococcal conjugate vaccine-naïve (PCV-naive) infants as established by medical history and clinical assessment before entering into the study. PCV-naïve infants are those who have not been previously vaccinated with any licensed or investigational pneumococcal vaccine.
2. Infants between 6-8 weeks of age (42-56 days, both days inclusive) of either gender, at the time of first dose of vaccination.
3. Healthy Infants with weight ≥ 3300 gms at the time of screening.
4. Subjects’ parent(s)/ LAR(s) who, in the opinion of the investigator, can and will comply, with the requirements of the protocol (e.g. completion of the diary cards, return for follow-up visits, with access to a consistent means of telephone contact, either residential landline or mobile).
5. Subject’s parent(s)/LAR(s) willing to provide written or thumb printed informed consent (including audio visual recording of consent process) prior to performing any study specific procedure
6. Infants with a minimal vaccination status for their age at the time of enrolment (“minimal” defined as single dose of BCG, Hepatitis B &/or Polio vaccine at the time of enrolment).

**Exclusion Criteria**

Subjects were excluded from the study based on the following exclusion criteria:

1. Child in care, defined as a child who has been placed under the control or protection of an agency, organisation, institution or entity by the courts, the government or a government body, acting in accordance with powers conferred on them by law or regulation. The definition of a child in care can include a child cared for by foster parents or living in a care home or institution, provided that the arrangement falls within the definition above. The definition of a child in care does not include a child who is adopted or has an appointed legal guardian.
2. Evidence of previous Streptococcus pneumoniae infection or pneumococcal vaccination.
3. Use of any investigational or non-registered product (drug or vaccine) during the period starting 30 days before the administration of study vaccine (Day -29 to Day 0), or planned use during the study period other than the study vaccine.
4. Any medical condition that in the judgment of the investigator would make intramuscular injection unsafe (eg., coagulation abnormalities).
5. Chronic administration (defined as more than 14 days in total) of immunosuppressants or other immune-modifying drugs or any blood products during the period starting 30 days prior to the proposed first vaccine dose or planned administration of the same during the study period.
6. Concurrently participating in another clinical study, at any time during the study period, in which the subject has been or will be exposed to an investigational or a non-investigational vaccine/product (pharmaceutical product or device).
7. Any confirmed or suspected immunosuppressive or immunodeficient condition, based on medical history and physical examination (no laboratory testing required).
8. Family history of congenital or hereditary immunodeficiency.
9. History of allergic disease or history of a serious reaction to any prior vaccination or known hypersensitivity likely to be exacerbated by any component of the study vaccines.
10. History of any neurological disorders, meningitis or seizures.
11. Infant who has had a sibling die of sudden infant death syndrome (SIDS) or die suddenly and without apparent other cause or preceding illness in the first year of life.
12. Infant is a direct descendant (child or grand-child) of any person employed by the Sponsor, the Contract Research Organization (CRO) or the Study Site (including the PI and study site personnel).
13. Acute disease and/or fever at the time of vaccination.
14. o Fever is defined as the endogenous elevation of at least one measured body temperature of ≥ 38◦C (≥ 100.4◦F).
15. Acute or chronic, clinically significant pulmonary, cardiovascular, hepatic or renal functional abnormality, as determined by physical examination and Principal investigator judgement.

**List of Solicited AEs:**

**Solicited Local AEs:** Pain at injection site, Redness at injection site, Swelling at injection site and Induration at injection site

The following general AEs are known to occur for similar marketed products (Prevenar13®):

**Solicited general AEs**

Irritability (very common)

Somnolence; Drowsiness/increased sleep; restless sleep/decreased sleep (very common)

Fever-mild (very common)

Seizures (including febrile seizures) (uncommon)

Hypotonic-hyporesponsive episode (rare)

Diarrhoea (common)

Vomiting (common)

Rash-Urticaria like (common)

Hypersensitivity reaction including face edema, dyspnea, bronchospasm (rare)

**List of study Investigators**

| **S. No** | **Site Code** | **Name of the Investigator** | **Institutional Affiliations And Site Locations** |
| --- | --- | --- | --- |
| 1. | **A** | Dr. G. Bala Kishor | Dept. of Paediatrics St. Theresa’s Hospital, Sanathnagar, Hyderabad – 500018 Telangana, India |
| 2. | **C** | Dr. S. Prashanth | Dept. of Paediatrics, Cheluvamba Hospital  Mysore Medical College & Research Institute, Irwin Road, Mysore – 570001, Karnataka, India |
| 3. | **D** | Dr. Ramchandra Keshav Dhongade | Dept. of Paediatrics, Sant Dnyaneshwar Medical Education & Research Centre, Opp.: Vijay Cinema Laxmi Road, Pune – 411030 Maharashtra, India. |
| 4. | **I** | Dr. Mane Sushant Satish | Grant Govt. Medical College and Sir J.J Group of Hospitals Byculla, Mumbai - 400008 Maharashtra, India. |
| 5. | **J** | Dr. M.D. Ravi | Dept. of Pediatrics JSS Hospital,, Mahatma Gandhi Road, Mysuru - 570004, Karnataka, India. |
| 6. | **K** | Dr. Savita Verma | Dept. of Paediatrics, PT. B D Sharma Post Graduate Institute of Medical Sciences & Hospital  Rohtak - 124001, Haryana, India. |
| 7. | **L** | Dr. V.N. Tripathi | Dept. of Paediatrics, Prakhar Hospital  8/219, Arya Nagar, Kanpur - 208002 Uttar Pradesh, India. |
| 8. | **N** | Dr. P. Venugopal | King George Hospital Collectorate Junction,  Maharanipeta, Visakhapatnam – 530002 Andhra Pradesh, India. |
| 9. | **O** | Dr. N.S. Mahantashetti | Dept. of Paediatrics  KLEs Prabhakar Kore Hospital & Medical Research Centre, Nehru Nagar  Belagavi - 590010, Karnataka, India |
| 10. | **P** | Dr. Jai Prakash Narayan | JLN Medical College, Kala Bagh, Ajmer 305001, Rajasthan, India |
| 11. | **R** | Dr. Abhishek T. Chavan | Jeevan Rekha Hospital, Dr. B R Ambedkar Road, Belagavi- 590002, Karnataka, India |
| 12. | **S** | Dr. Srimukhi Anumolu | Vignesh Women & Children Hospital, Prajasakti Nagar, Vijayawada, Andhra Pradesh, India |

**2. Institutional Ethics Committees (IECs) and approvals**

| S. No. | Site Code | Site Name | PI Name & Address | EC Approval Date |
| --- | --- | --- | --- | --- |
| 1 | A | St.Theresas Hospital | Dr. G. Bala Kishor    St.Theresas Hospital (STH), 1st Floor, Room No. 05, Erragadda Main Road, Czech Colony Sanath Nagar, Hyderabad 500038, Telangana, India | 27.01.20 |
| 2 | C | Cheluvamba | Dr. S. Prashanth Cheluvamba Hospital, Irwin Rd, Devraj Mohalla, Mysure-570001, Karnataka , India | 25.01.20 |
| 3 | D | Sant Dnyaneshwar | Dr. Ramchandra Keshav Dhongade Sant Dnyaneshwar Medical Education & Research Centre, Pune | 01.07.21 |
| 4 | I | Grant Medical College | Dr. Mane Sushant Satish Grant Medical College & Sir J.J Hospital, J J Marg, Nagpada, Mumbai Central, Mumbai 400008, Maharashtra, India | 05.06.21 |
| 5 | J | JSS Hospital | Dr. M. D. Ravi JSS Hospital, Mahatma Gandhi Road, Fort Mohalla, Mysuru-570004, Karnataka, India | 07.08.20 |
| 6 | K | Pandit Bhagwat | Dr. Savita Verma Pandit Bhagwat Dayal Sharma Post Graduate Institute of Medical Sciences & Hospital, Rothak, Haryana | 23.01.20 |
| 7 | L | Prakhar Hospital | Dr. VN Tripathi Prakhar Hospital, 8/219, Khalasi Line, Arya Nagar, Kanpur 208002, Uttar Pradesh, India | 27.01.20 |
| 8 | N | KGH | Dr. P Venugopal Department of Paediatric, King George Hospital Collectorate Junction, Maharanipeta Visakhapatnam – 530002 .Andhra Pradesh, India. | 27.01.20 |
| 9 | O | KLE | Dr. NS Mahantashetti KLES Dr. Prabhakar Kore Hospital & Medical Research Centre, J N Medical College,Nehru Nagar, Belagavi – 590010,Karnataka, India | 06.02.20 |
| 10 | P | JLN Medical college | Dr. Jai Prakash Narayan JLN Medical College , Kala Bagh, Ajmer 305001, Rajasthan, India | 18.10.21 |
| 11 | R | Jeevan Rekha | Dr. Abhishek T. Chavan Jeevan Rekha Hospital, Dr. B R Ambedkar Road,Belagavi- 590002, Karnataka, India | 01.10.21 |
| 12 | S | Vignesh | Dr. Srimukhi Anumolu Vignesh Women & Children Hospital, Prajasakti Nagar, Vijayawada, Andhra Pradesh, India | 09.10.21 |
